# Depressive Symptoms and its Associated Factors among Health Care Workers in Saint Paul’s Hospital Millennium Medical College, Ethiopia

**DOI:** 10.1101/2024.03.05.24303845

**Authors:** Melese Bahiru Tesema, Berhanu Teshome Woldeamanue, Eyoel Berhane Mekonen, Kidest Getu Melese

## Abstract

**Background:** Depression is a mental disorder that presents with a depressed mood, loss of pleasure or interest, decrease in energy, feeling of guilt or low self-worth, disturbed sleep, loss of appetite, and poor-concentration. Healthcare professionals are more susceptible to depression because they face higher amounts of professional stress in their job and academic lives. However, there is limited knowledge regarding health professionals’ level of depressive disorder in Ethiopia. This study aimed to assess the prevalence of depression and associated factors among health professionals, at Saint Paul’s Hospital Millennium Medical College, Addis Ababa, Ethiopia.

**Methods:** We conducted an institution-based cross-sectional study among 439 randomly selected health care worker using interviewer-administered patient health questionnaire-9 from April to May 2023. Ordinal logistic regression was performed to identify predictors of depressive disorder.

**Result:** The overall prevalence of depressive symptoms among health care workers was 56.5% (95% CI (51.8%, 61.1%)). There were reports of mild (35%), moderate (13%) and severe (9%) depression, respectively. Marital status of being single (AOR=7.78, 95%CI: 1.123, 49.01), history of childhood abuse (AOR=2.57, 95%CI:1.49, 4.42), history of suicidal attempt (AOR=2.66, 95%CI:1.25,5.67), having history of stressful life event (AOR=1.527, 95%CI: 1.02,2.3), back pain over the past 30 days (AOR=2, 95%CI: 1.30,3.11), working for more than 8 hours (AOR=3.03, 95%CI: 1.12,8.24), and having experience of 5-10 year (AOR=4, 95%CI: 1.05,15.27) and 10-15 years (AOR= 4.24, 95%CI: 1.08,16.58) and poor social support (AOR= 2.09, 95%CI: 1.09,3.99) were statistically associated with increased level of depressive disorder.

**Conclusion:** Healthcare professionals’ higher rate of depression was due to the higher work load, childhood abuse, history of stressful life, back pain, and poor social support. Thus, the hospital or ministry of health should give special attention to health care workers who has high work load, childhood abuse, history of stressful life, back pain, and poor social support to reduce the burden of depressive disorder among health professionals. Furthermore, early depression screening and treatment in hospitals is needed.

## Background

Depressive disorder (DD) are characterized by sadness, lack of interest or enjoyment, feelings of guilt or low self-worth, disturbed sleep or appetite, severe weight loss, fatigue, poor focus, psychomotor agitation or retardation, and recurring thoughts of death (suicide) (1). Key signs of depression include low mood and a lack of interest (2). Individuals, who have major depressive disorder (MDD) encounter more physical illness and pain, along with decreased physical and social role functioning (3). DD has consistently been among the top ten leading causes of all years lived with disability (YLDs) worldwide and the second leading cause of global YLDs, accounting for 5.6% of all YLDs in 2019 (4). Individuals with a diagnosis of MDD had 40–60% higher probability than the general population to die at younger age (5).

In contrast to the general population, various studies have documented health care workers (HCWs) are at greater risk of exposure to potential risk of depression because of the demanding nature of their jobs (6–8). In developed countries the prevalence of depression ranges from 21.53% to 32.77% among HCWs(9–13). A systematic review and meta-analysis study done using 57 cross-sectional studies from 2019 to 2020 reported 24% pooled prevalence rate of depression among HCWs and 43% among frontline HCWs (14). A similar study done in 2020, in Wuhan, China, among 994 doctors and nurses working in a high-risk environment using instrument Patient Health Quastionnare-9 (PHQ-9) found, 36% of medical staff shows sub-threshold level of depression, 34.4% had mild depression, 22.45% had moderate depression and 6.2% had severe depression (15). Further, in Nigeria, the prevalence of DD was 17.3% among resident doctors and 1.3% among non-resident doctors (6). A similar study done in a primary health care center in Egypt, using PHQ-9 among Doctors and Nurses in teaching hospitals showed the prevalence of depression was 71.4% (7). According to Shah J et al, one-half of HCWs (53.6%) in Kenya tested positive for depression, with the majority suffering from moderate depression (8). Further studies undertaken among HCWs in Botswana, Uganda, and Sudan using the DASS-21, found 21%, 40.3% and 75% prevalence of depression, respectively (16–18).

In Ethiopia, a systematic review and meta-analysis study reported a pooled prevalence rate of depression among HCWs was 11% (19). In eastern Ethiopia, the prevalence of minimal, mild, moderate, and severe depressive symptoms were 27.9%, 24.1%, 9.4%, and 1.1%, respectively (20). Another study in central Ethiopia (21), southern Ethiopia (22), and Gondar (23) reported highest prevalence of DD, 60.3%, 50.1%, and 42.7% respectively.

The common risk factors associated with depressive symptoms are the clinical specialty, presence of chronic illness, substance use, history of psychiatric disorder, insufficient social support, family history of mental illness, being unmarried, low income, long working hours, being female and working at night time (4).

Although similar studies have been done in Ethiopia, the focus of this study is on the prevalence of DD and factors related to it among healthcare professionals. This study therefore attempts to evaluate the prevalence of DD and factors related to it among HCWs. Determining the frequency and contributing variables of depressive symptoms also provides knowledge, validates the results of other studies, and may serve as the basis for developing novel intervention strategies. Therefore, this study aims to assess the prevalence of depressive disorder and its associated factor among health care workers in Saint Paul’s Hospital Millennium Medical College, Addis Ababa, Ethiopia.

## Methods

### Study Design and Setting

Institution based cross-sectional study was conducted from April 3/ 2023 to May 10/ 2023 at Saint Paul’s Hospital Millennium Medical College, Addis Ababa, Ethiopia. Saint Paul’s hospital millennium medical college, as it is known today, was established through a decree of the council of ministers in 2010, although the medical school was opened in 2007 and the hospital was established in 1968 by the late Emperor Haileselassie I (24). It is governed by board under federal ministry of health. The hospital provides preventive, curative and rehabilitative care for patient coming from health center by referral system. The inpatient capacity of the hospital is more than 700 beds, and the college sees an average of 1200 emergency and outpatient clients daily (24). Since the hospital has high patient flow both in outpatient and inpatient setup; HCWs working there become susceptible to work related depression.

### Sample Size and study population

We used double population proportion formula to calculate sample size by using state calc of Epi-info. By using 95% CI, 80% power, taking ratio of unexposed to exposed, odd ratio and proportion of respective variable of exposure and adding 10% non-response rate the largest sample size of 439 was taken as final sample size (Table 1). The study participants were selected through stratified random sampling method by taking profession as their strata. The sample size was allocated proportional to size of a strata and finally samples were selected using simple random sampling technique within a strata (Fig 1. Proportional Allocation to Study the Prevalence of Depression and Associated Factors Among Health Care Professionals Working at SPHMMC, Addis Ababa, Ethiopia). (SPHMMC=Saint Paul’s Hospital Millenium medical college, GP=General Practitioner, Lab. Professional’s=Laboratory Professional’s)

**Table 1.** Assumption used to calculate sample size by double population proportion using stat-calc of Epi-info.

| S.no | Variable as exposures | Assumptions |  |  |  |  | Sample size | Final sample size by adding 10% non-response rate |
| --- | --- | --- | --- | --- | --- | --- | --- | --- |
|  |  | OR | P | Ratio | Power | CI |  |  |
| 1 | Sex (Female) | 1.94 | 24.5% | 1.83 | 80% | 95% | 395 | 435 |
| 2 | Marital status (single) | 2.16 | 22.1% | 0.70 | 80% | 95% | 296 | 326 |
| 3 | Family history of mental illness | 7.31 | 24.4% | 13 | 80% | 95% | 146 | 161 |
| 4 | Smoker | 2.67 | 22.2% | 3 | 80% | 95% | 222 | 244 |
| 5 | Workload (Hours) | 1.997 | 69 | 1.16 | 80% | 95% | 399 | 439 |

**Fig 1.**
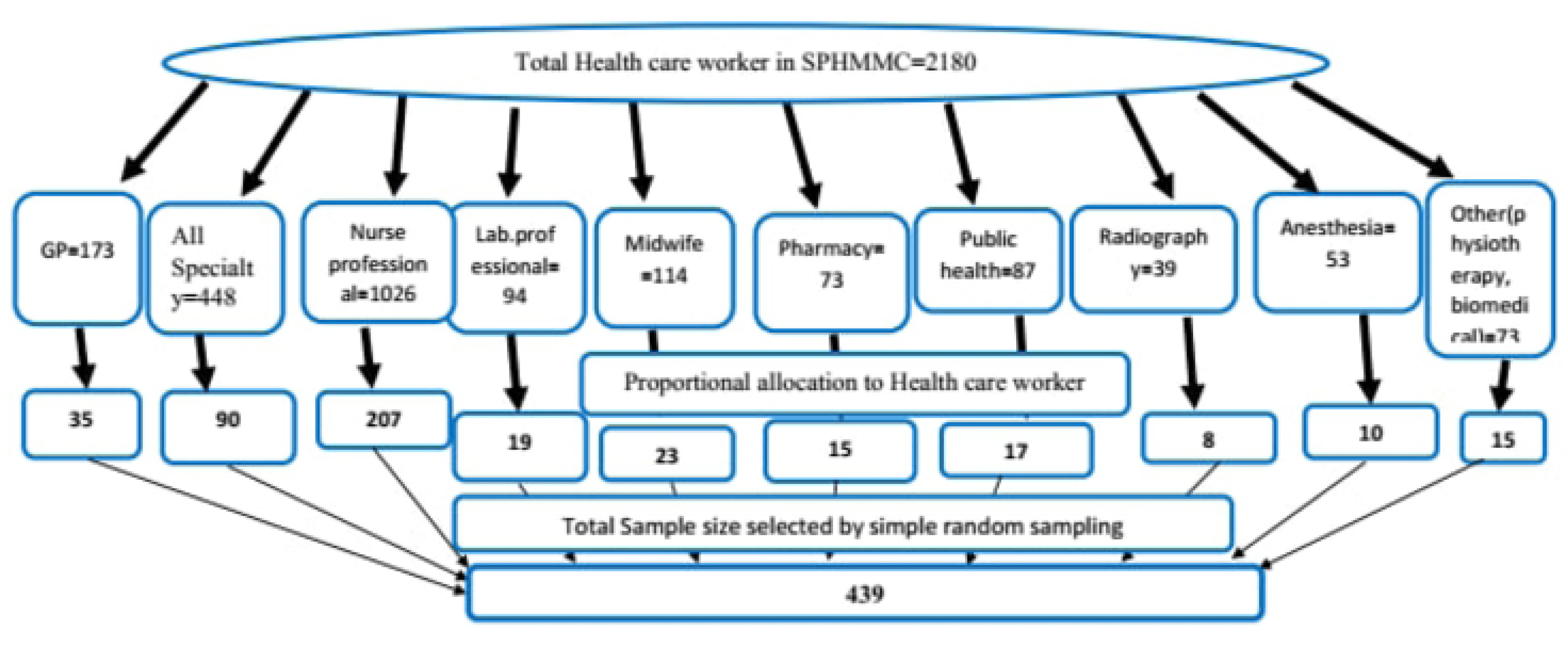

### Measurement of Study Variable

Depressive Disorder was the outcome variable of this study and the exposure variables were sociodemographic variables (age, sex, marital status, religion, level of education and monthly income), and psycho-social factors (history of childhood abuse, stressful life event, history of suicidal attempt, presence of family history of mental illness). Further, physical health status (history of injury, presence of disability, headache over past 30 days, back pain over the past 30 days, fever over past 30 days, medical problem, smoking history and social support), health professional status and job satisfaction level (field of study, working hour, work experience, work environment, working department and job satisfaction) were used. PHQ-9 was used to assess DD with nine items of major depressive disorder symptoms from DSM-IV criteria. PHQ-9 was validated previously with sensitivity of 86% and specificity of 67% in Ethiopian population (25). Total score of nine items of PHQ-9 ranging from 0 to 27, having each item response rated as ‘’0’’ (not at all) to ‘’3’’ (nearly every day). No depressive symptom, mild, moderate, moderately severe and severe depressive symptoms were defined by tool as total scores of 0-4,5-9,10-14, 15-19 and 20 and above, respectively (26). For this study scores less than 5 was taken as no depressive symptom, 5-9 (mild depressive symptom), 10-14 (moderate depressive symptom), 15 and above as severe depressive symptom. To assess social support Oslo-3-item social support scale was used and those with score of 3-8 was classified as poor social support, 9-11 as moderate social support and 12-14 as strong social support (27).

### Data Collection

The data was collected using interviewer administrated Questionnaire. The recruitment period for this study was from April 03/2023 to May 10/2023. Two trained health professionals were recruited to collect data and two of the authors supervised the data collection. Before the data collection pre-test with 5% of health professionals was conducted at Ras Desta Damtew Referral Hospital located nearby Saint Pauls Hospital Millennium Medical College. Completeness and consistency of items was ensured to minimize systematic error.

### Statistical Analysis

The data was checked and cleaned in Excel form and then exported to Statistical Package for the Social Science version 26 for coding and analysis. A descriptive summary was used to present the study results. Both bivariable and multivariable ordinal logistic regression model was fitted to determine the association between the dependent and the independent variables. Only factors that are significantly associated during bivariable analysis with a p-value less than 0.25 were entered into the multivariable analysis. In this regard, the Likelihood ratio test of goodness-of-fit test was used. Multi-collinearity was checked with the Variance inflation factor. Adjusted odds ratio with 95% CI, p-value <0.05 was reported.

## Result

### Descriptive Statistics

Out of 439 HCWs participated in the study, 241(55%) were males. The mean age of the respondents was 31.23 years with standard deviation of 6.23. Fifty percent of the respondents’ (n=220) were single, 206(46.9%) married, 9(2.1%) divorced. Concerning education level 13(3%) has college diploma, 236(53.8%) has Bachler degree, 63(14.4%) has master degree, 127(28.9%) has doctorate degree. Further, regarding religion, 292(66.5 %) are Orthodox, 93(21.2%) were protestant and 49(11.2%) were Muslim followers. Concerning monthly income 202(46 %) of the respondents had earned less than eight thousand (8000) Ethiopian Birr or 141.72 United State Dollar (Table 2).

**Table 2.** Distribution of Socio-Demographic Variables among HCWs of Saint Pauls’ Hospital Millennium Medical College, Addis Ababa, Ethiopia,2023 (n=439)

| **Variable** | **Category** | **% (n)** |
| --- | --- | --- |
| Sex | Male | 54.9 (241) |
|  | Female | 45.1 (198) |
| Age | 18-25 year | 10 (44) |
|  | 26-30 year | 49.4 (217) |
|  | 31-35 year | 20.3 (89) |
|  | 36-40 year | 13 (57) |
|  | 41-60 year | 7.3 (32) |
| Marital status | Single | 50.1(220) |
|  | Married | 46.9(206) |
|  | Divorced | 3(13) |
| Level of education | College Diploma | 3(13) |
|  | Bachelor Degree | 53.8(236) |
|  | Master Degree | 14.4(63) |
|  | Doctorate Degree | 28.9(127) |
| Religion | Orthodox | 66.5(292) |
|  | Protestant | 21.2(93) |
|  | Muslim | 11.2(49) |
|  | Other | 1.1(5) |
| Monthly income | <8000birr | 46(202) |
|  | 8001-10,000 birr | 25.7(113) |
|  | 10001-14999birr | 18.9(83) |
|  | >15000birr | 9.3(41) |

As the psychosocial factors were concerned twenty percent of the study participants faced childhood abuse, 51(11.6%) had a history of mental illness in their family, 36(8.2%) had a suicidal attempt and 212(48.3%) had a history of stressful life situations. Among those who sustain childhood abuse, 34(38.2%) are sustained by physical abuse, 26(29.2%) by verbal abuse and 22(24.7%) by emotional abuse. Furthermore, among those who sustained stressful life events 25(45.5%) were due to the loss of a close family member or loved one, and 17(30.9%) were due to the loss of money or materials (Table 3).

**Table 3.** Distribution of Psychosocial factor among HCWs of Saint Pauls’ Hospital Millennium Medical College,2023 (n=439)

| **Variable** | **Category** | **% (n)** |
| --- | --- | --- |
| History of childhood abuse | Yes | 20.5(90) |
|  | No | 79.5(349) |
| Presence of family history of mental illness | Yes | 11.6(51) |
|  | No | 88.4(388) |
| History of suicidal attempt | Yes | 8.2(36) |
|  | No | 91.8(403) |
| History of stress full life event | Yes | 48.3(212) |
|  | No | 51.7(227) |

Regarding the physical status and social support, 45(10.3%) had sustained injury, 11(2.5%) had a disability, and 42(9.6%) had a medical problem. Additionally, 146(33.3%) of study participants had headaches, 151(34.4%) had back pain, and 47(10.7%) had fever over the past 30 days respectively. Further, 31(7.1%) of the study participants had smoking history. According to the Oslo 3-item social support scale, those who have poor, moderate, and strong social support constitute 156(35.5%), 223(50.8%) and 60(13.7%) of the study participants, respectively (Table 4).

**Table 4.** Distribution of Physical health status and social support among HCWs of Saint Pauls’ Hospital Millennium Medical College,2023(n=439)

| Variable | Category | %(n) |
| --- | --- | --- |
| History of Injury | Yes | 10.3(45) |
|  | No | 89.7(394) |
| Presence of disability | Yes | 2.5(11) |
|  | No | 97.5 (428) |
| Presence of head ache over the past 30 days | Yes | 33.3(146) |
|  | No | 66.7(293) |
| Presence of back pain over the past 30 days | Yes | 34.4(151) |
|  | No | 65.6(288) |
| Presence of fever over the past 30 days | Yes | 10.7(47) |
|  | No | 89.3(392) |
| Presence of medical problem | Yes | 9.6(42) |
|  | No | 90.4(397) |
| Smoking history | Yes | 7.1(31) |
|  | No | 92.9(408) |
| Social support | Poor | 35.5(156) |
|  | Moderate | 50.8(223) |
|  | Strong | 13.7(60) |

Almost half of the study participants were nurses. There were 90(20.5%) medical specialties including residency, and 35(8%) general practitioners. In terms of other professions, 23(5.2%) are midwives, 19(4.3%) are medical laboratories, 17(3.9%) are public health, 15(3.4%) are pharmacists, 10(2.3%) are anesthesiologists, 8(1.8%) are radiologists, and 15(3.4%) are others such as Psychiatry professionals, Phlebotomist, Biomedical engineer, and Biomedical technician.

Regarding the years of work experience 191(43.5%) of study participants had 2-5 years of experience, and 146(33.3%) had 10-15 years of experience. Regarding the working department and environment, 88(20%) work in the medical department, 79(18.3%) work in the surgical department, 13 (3%) in the ophthalmic and 11(2.5%) in the ear, nose and throat department. Further, 95(21.6%) of respondents’ working environment is emergency, and 190(43.3%) work as inpatient. Moreover, regarding working hours and level of job satisfaction, 278(63.3%) work more than 8 hours and 207(47%) of respondents currently dissatisfied with their job (Table 5).

**Table 5.**
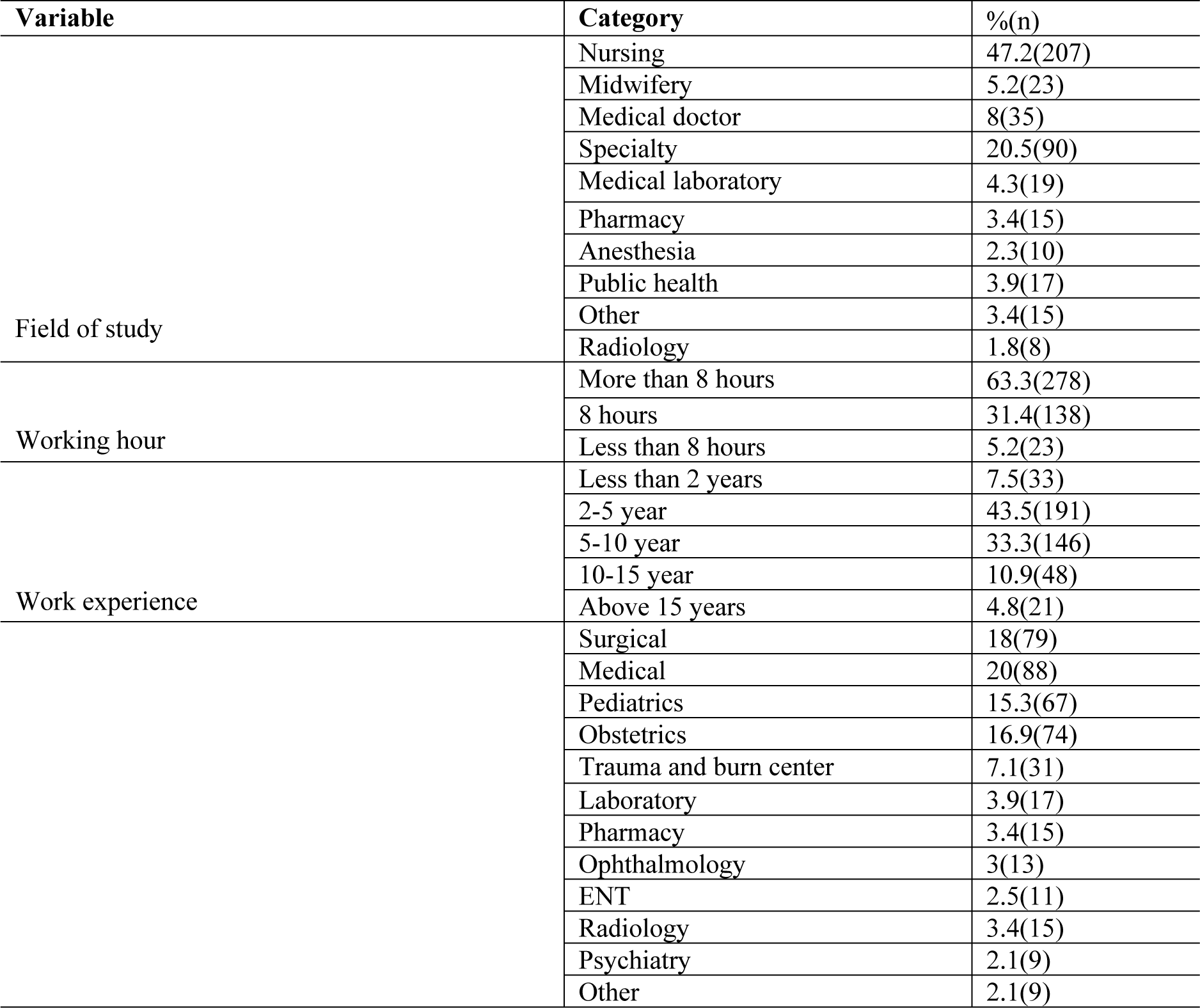
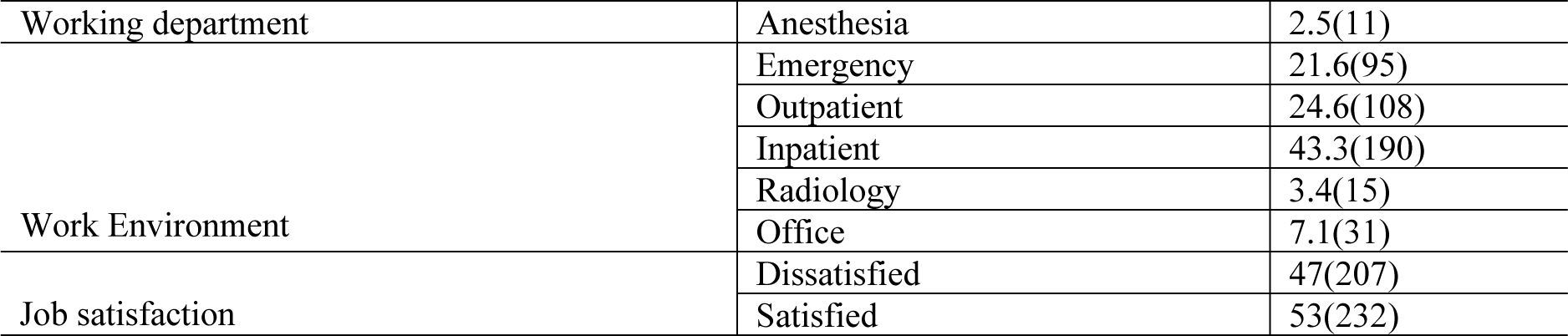
Distribution of Professional factor, Job Satisfaction and work area factor among HCWs of Saint Pauls’ Hospital Millenium Medical college,2023(n=439)

### Prevalence of Depression among Health Care Worker at Saint Paul Hospital Millennium Medical College

Based on the cut of point ≥5 for cases on PHQ-9, the prevalence of depression among HCWs in SPHMMC was found 248 (56.5% (95%CI: 35.3%, 77.7%)), of which 138 (55.6%) were male. Concerning the severity of symptoms, 41(9.3%) have severe depressive symptoms, 55(12.5%) have moderate depressive symptoms, and 152(34.6%) have mild depressive symptoms (Fig 2. Prevalence of Depression by Severity Among HCWs, in Saint Paul’s Hospital Millennium Medical College, Addis Ababa, Ethiopia, 2023).

**Fig 2.**
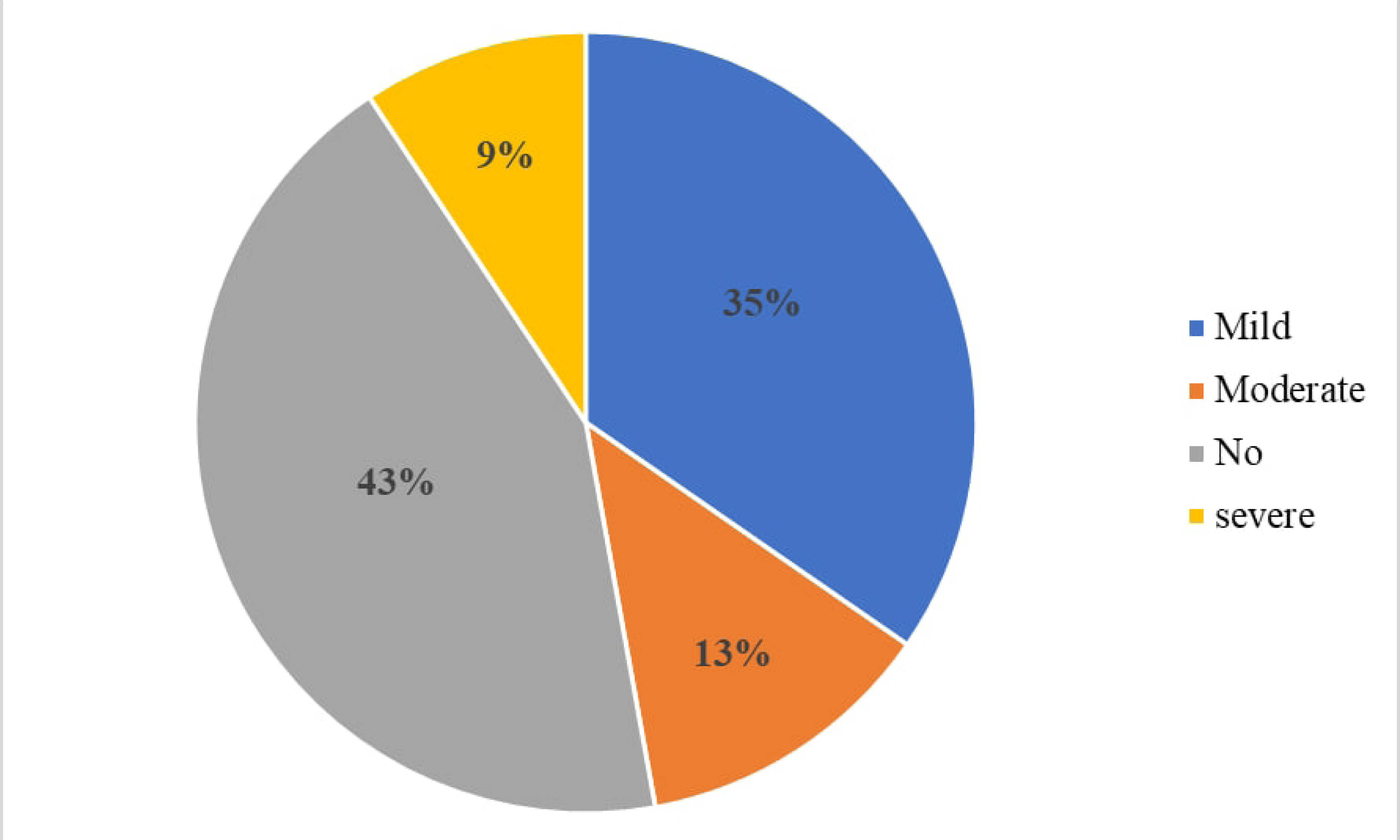

### Factors associated with Depression among Health Care Worker at Saint Pauls’ Hospital Millennium Medical College

In model fitting, we started with all candidate predictors significant at 25% in the bivariate analysis and then employed backward elimination methods to reach the final model. The overall good fit of the final model was assessed using, the Likelihood ratio test of goodness-of-fit test (Chi-square=130.753) with p-value <0.001 indicating good fit of the model. Having fitted the proportional odds model, the proportionality assumption for the fitted proportional odds model was checked using the likelihood ratio test, which confirms no violation of this assumption (Chi-square = 28, p-value =0.999). Multi-collinearity was checked with Variance inflation factor. There is no multicollinearity problem found.

In the multivariable analysis, being single (AOR=7.78, 95% CI (1.23, 49.011) was found to be highly associated with increased odds of depression than divorced state. Further, sustained childhood abuse (AOR=2.57, 95% CI (1.49, 4.42)), history of suicidal attempt (AOR=2.66, 95% CI (1.25, 5.67)), and history of stressful life event (AOR=1.527, 95% CI (1.02, 2.3)) were associated with a greater odd of depression, compared to their counterpart did not sustain childhood abuse, no history of suicidal attempt and no history of stressful life event, respectively.

Furthermore, presence of back pain over the past 30 days (AOR=2, 95% CI (1.3, 3.11)), working more than 8 hour (AOR=3.03, 95% CI (1.119, 8.245)), work experience of 5-10 year (AOR=4, 95% CI (1.05, 15.27)), work experience of 10-15 year (AOR=4.24, 95% CI (1.08, 16.58)), poor social support (AOR=2.09(1.09, 3.99)) were associated with increased odds of higher depression level, than their counter part (Table 6).

**Table 6:**
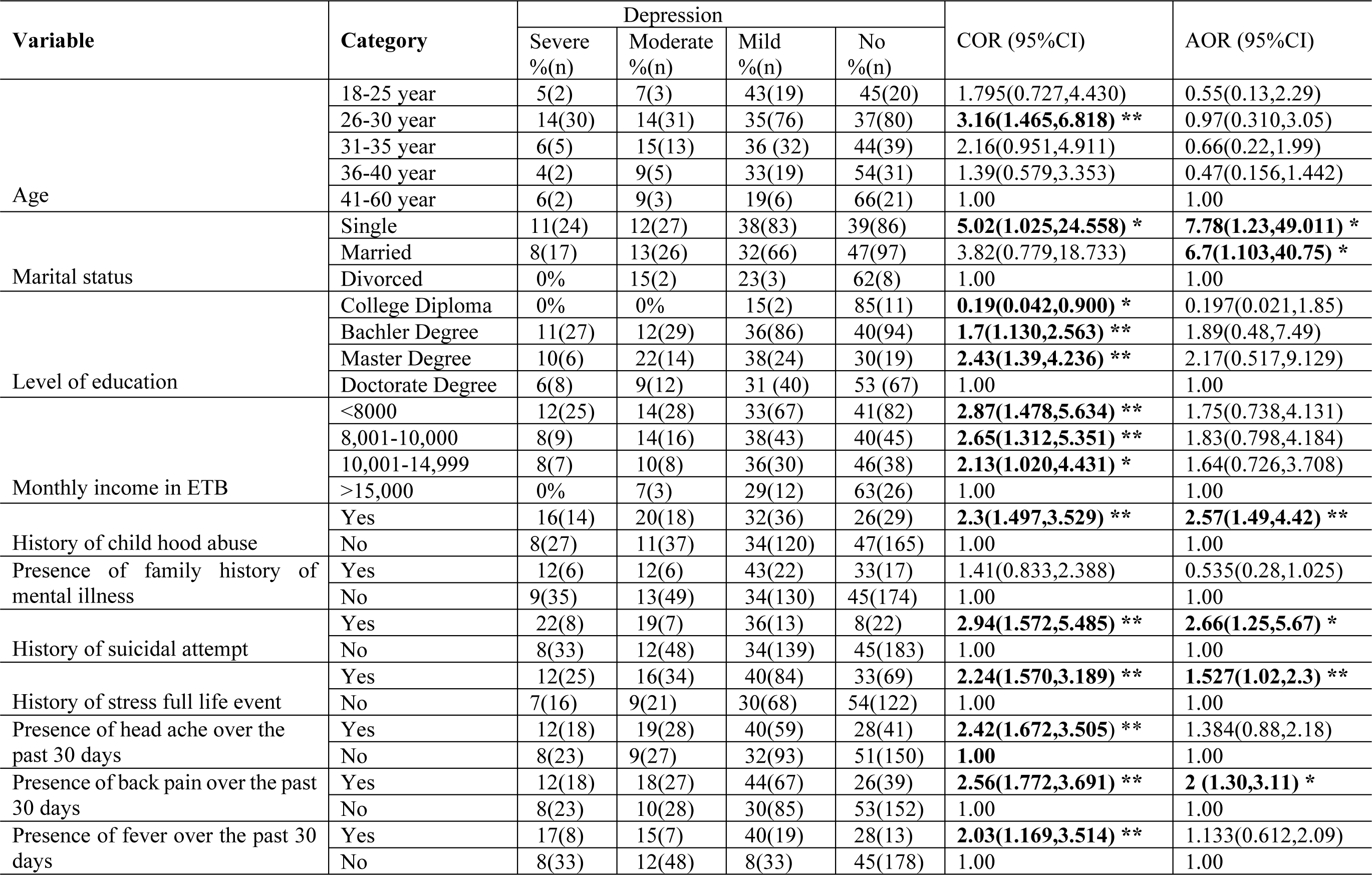
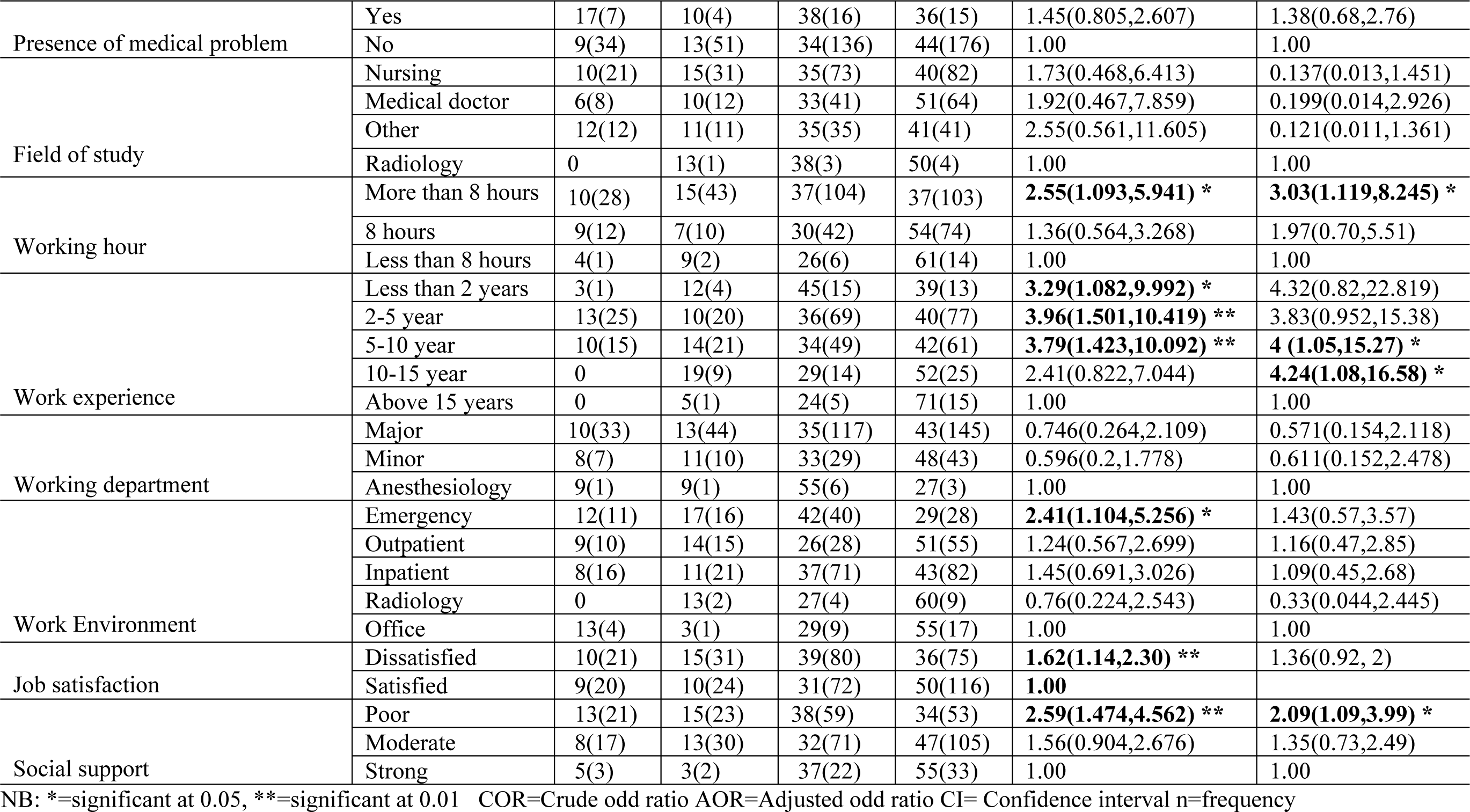
Multivariable Analysis of Different Variable and Depression among HCWs of Saint Pauls’ Hospital Millennium Medical College Addis Ababa, Ethiopia,2023(n=439)

| Variable | Category | Depression |  |  |  | COR (95%CI) | AOR (95%CI) |
| --- | --- | --- | --- | --- | --- | --- | --- |
|  |  | Severe<br>%(n) | Moderate<br>%(n) | Mild<br>%(n) | No<br>%(n) |  |  |
| Age | 18-25 year | 5(2) | 7(3) | 43(19) | 45(20) | 1.795(0.727,4.430) | 0.55(0.13,2.29) |
|  | 26-30 year | 14(30) | 14(31) | 35(76) | 37(80) | **3.16(1.465,6.818) \*\*** | 0.97(0.310,3.05) |
|  | 31-35 year | 6(5) | 15(13) | 36 (32) | 44(39) | 2.16(0.951,4.911) | 0.66(0.22,1.99) |
|  | 36-40 year | 4(2) | 9(5) | 33(19) | 54(31) | 1.39(0.579,3.353) | 0.47(0.156,1.442) |
|  | 41-60 year | 6(2) | 9(3) | 19(6) | 66(21) | 1.00 | 1.00 |
| Marital status | Single | 11(24) | 12(27) | 38(83) | 39(86) | **5.02(1.025,24.558) \*** | **7.78(1.23,49.011) \*** |
|  | Married | 8(17) | 13(26) | 32(66) | 47(97) | 3.82(0.779,18.733) | **6.7(1.103,40.75) \*** |
|  | Divorced | 0% | 15(2) | 23(3) | 62(8) | 1.00 | 1.00 |
| Level of education | College Diploma | 0% | 0% | 15(2) | 85(11) | **0.19(0.042,0.900) \*** | 0.197(0.021,1.85) |
|  | Bachler Degree | 11(27) | 12(29) | 36(86) | 40(94) | **1.7(1.130,2.563) \*\*** | 1.89(0.48,7.49) |
|  | Master Degree | 10(6) | 22(14) | 38(24) | 30(19) | **2.43(1.39,4.236) \*\*** | 2.17(0.517,9.129) |
|  | Doctorate Degree | 6(8) | 9(12) | 31 (40) | 53 (67) | 1.00 | 1.00 |
| Monthly income in ETB | <8000 | 12(25) | 14(28) | 33(67) | 41(82) | **2.87(1.478,5.634) \*\*** | 1.75(0.738,4.131) |
|  | 8,001-10,000 | 8(9) | 14(16) | 38(43) | 40(45) | **2.65(1.312,5.351) \*\*** | 1.83(0.798,4.184) |
|  | 10,001-14,999 | 8(7) | 10(8) | 36(30) | 46(38) | **2.13(1.020,4.431) \*** | 1.64(0.726,3.708) |
|  | >15,000 | 0% | 7(3) | 29(12) | 63(26) | 1.00 | 1.00 |
| History of child hood abuse | Yes | 16(14) | 20(18) | 32(36) | 26(29) | **2.3(1.497,3.529) \*\*** | **2.57(1.49,4.42) \*\*** |
|  | No | 8(27) | 11(37) | 34(120) | 47(165) | 1.00 | 1.00 |
| Presence of family history of mental illness | Yes | 12(6) | 12(6) | 43(22) | 33(17) | 1.41(0.833,2.388) | 0.535(0.28,1.025) |
|  | No | 9(35) | 13(49) | 34(130) | 45(174) | 1.00 | 1.00 |
| History of suicidal attempt | Yes | 22(8) | 19(7) | 36(13) | 8(22) | **2.94(1.572,5.485) \*\*** | **2.66(1.25,5.67) \*** |
|  | No | 8(33) | 12(48) | 34(139) | 45(183) | 1.00 | 1.00 |
| History of stress full life event | Yes | 12(25) | 16(34) | 40(84) | 33(69) | **2.24(1.570,3.189) \*\*** | **1.527(1.02,2.3) \*\*** |
|  | No | 7(16) | 9(21) | 30(68) | 54(122) | 1.00 | 1.00 |
| Presence of head ache over the past 30 days | Yes | 12(18) | 19(28) | 40(59) | 28(41) | **2.42(1.672,3.505) \*\*** | 1.384(0.88,2.18) |
|  | No | 8(23) | 9(27) | 32(93) | 51(150) | **1.00** | 1.00 |
| Presence of back pain over the past 30 days | Yes | 12(18) | 18(27) | 44(67) | 26(39) | **2.56(1.772,3.691) \*\*** | **2 (1.30,3.11) \*** |
|  | No | 8(23) | 10(28) | 30(85) | 53(152) | 1.00 | 1.00 |
| Presence of fever over the past 30 days | Yes | 17(8) | 15(7) | 40(19) | 28(13) | **2.03(1.169,3.514) \*\*** | 1.133(0.612,2.09) |
|  | No | 8(33) | 12(48) | 8(33) | 45(178) | 1.00 | 1.00 |
| Presence of medical problem | Yes | 17(7) | 10(4) | 38(16) | 36(15) | 1.45(0.805,2.607) | 1.38(0.68,2.76) |
|  | No | 9(34) | 13(51) | 34(136) | 44(176) | 1.00 | 1.00 |
| Field of study | Nursing | 10(21) | 15(31) | 35(73) | 40(82) | 1.73(0.468,6.413) | 0.137(0.013,1.451) |
|  | Medical doctor | 6(8) | 10(12) | 33(41) | 51(64) | 1.92(0.467,7.859) | 0.199(0.014,2.926) |
|  | Other | 12(12) | 11(11) | 35(35) | 41(41) | 2.55(0.561,11.605) | 0.121(0.011,1.361) |
|  | Radiology | 0 | 13(1) | 38(3) | 50(4) | 1.00 | 1.00 |
| Working hour | More than 8 hours | 10(28) | 15(43) | 37(104) | 37(103) | **2.55(1.093,5.941) \*** | **3.03(1.119,8.245) \*** |
|  | 8 hours | 9(12) | 7(10) | 30(42) | 54(74) | 1.36(0.564,3.268) | 1.97(0.70,5.51) |
|  | Less than 8 hours | 4(1) | 9(2) | 26(6) | 61(14) | 1.00 | 1.00 |
| Work experience | Less than 2 years | 3(1) | 12(4) | 45(15) | 39(13) | **3.29(1.082,9.992) \*** | 4.32(0.82,22.819) |
|  | 2-5 year | 13(25) | 10(20) | 36(69) | 40(77) | **3.96(1.501,10.419) \*\*** | 3.83(0.952,15.38) |
|  | 5-10 year | 10(15) | 14(21) | 34(49) | 42(61) | **3.79(1.423,10.092) \*\*** | **4 (1.05,15.27) \*** |
|  | 10-15 year | 0 | 19(9) | 29(14) | 52(25) | 2.41(0.822,7.044) | **4.24(1.08,16.58) \*** |
|  | Above 15 years | 0 | 5(1) | 24(5) | 71(15) | 1.00 | 1.00 |
| Working department | Major | 10(33) | 13(44) | 35(117) | 43(145) | 0.746(0.264,2.109) | 0.571(0.154,2.118) |
|  | Minor | 8(7) | 11(10) | 33(29) | 48(43) | 0.596(0.2,1.778) | 0.611(0.152,2.478) |
|  | Anesthesiology | 9(1) | 9(1) | 55(6) | 27(3) | 1.00 | 1.00 |
| Work Environment | Emergency | 12(11) | 17(16) | 42(40) | 29(28) | **2.41(1.104,5.256) \*** | 1.43(0.57,3.57) |
|  | Outpatient | 9(10) | 14(15) | 26(28) | 51(55) | 1.24(0.567,2.699) | 1.16(0.47,2.85) |
|  | Inpatient | 8(16) | 11(21) | 37(71) | 43(82) | 1.45(0.691,3.026) | 1.09(0.45,2.68) |
|  | Radiology | 0 | 13(2) | 27(4) | 60(9) | 0.76(0.224,2.543) | 0.33(0.044,2.445) |
|  | Office | 13(4) | 3(1) | 29(9) | 55(17) | 1.00 | 1.00 |
| Job satisfaction | Dissatisfied | 10(21) | 15(31) | 39(80) | 36(75) | **1.62(1.14,2.30) \*\*** | 1.36(0.92, 2) |
|  | Satisfied | 9(20) | 10(24) | 31(72) | 50(116) | **1.00** |  |
| Social support | Poor | 13(21) | 15(23) | 38(59) | 34(53) | **2.59(1.474,4.562) \*\*** | **2.09(1.09,3.99) \*** |
|  | Moderate | 8(17) | 13(30) | 32(71) | 47(105) | 1.56(0.904,2.676) | 1.35(0.73,2.49) |
|  | Strong | 5(3) | 3(2) | 37(22) | 55(33) | 1.00 | 1.00 |

NB: \*=significant at 0.05, \*\*=significant at 0.01 COR=Crude odd ratio AOR=Adjusted odd ratio CI= Confidence interval n=frequency

## Discussion

This study assessed the prevalence and the factors associated with DD among HCWs. The overall prevalence of DD in this study was 56.5% (95% CI (51.8%,61.1%)); while the prevalence of mild, moderate, and severe depression was 35%, 13%, and 9%, respectively. This is nearly agreed with research conducted in central Ethiopia 60.3% (21), China 57.2% (28), and Kenya 53.6%(8). However, the proportion of depressive symptoms, in our study was higher when compared to various studies conducted in Ethiopia; Eka kotebe Hospital 23.5% (29), Dessie comprehensive specialized hospital 27.8% (30), Gurage zonal public hospitals 25.8% (31), Saudi Arabia 43.9% (32), Southern China public hospital 38%(33), Southern Ethiopia 50.1% (22) and Gondar comprehensive specialized Hospital 42.7% (23). This discrepancy could be the result of different study designs and instruments; for example, the study conducted in Gondar and China utilized a DASS-21, whereas this study used a PHQ-9. Difference in sample size and professional mix may be other reason for discrepancy. Study done in China comprised of 3474 individuals, all of whom were nurses, and our study, included participants from a variety of health profession. Further, another potential explanation could be the variation in participants’ socioeconomic and demographic features within the communities. On the other hand, the findings from this study was lower compared to studies done in Eastern Ethiopia 66.4%(20), Baghdad 70.25%(34), Egypt 71.4%(35) and Sudan 75%(17). The discrepancy might be due to the inclusion criteria, sample size and instrument used. Moreover, variation in study participants, time variation, working environment and probably the difference in a medical setting might be the possible reason for the difference and those study was done during Era of COVID-19.

The factors associated with DD in this study were having a history of abuse as a child, having attempted suicide in the past, having back pain in the past, being single, working more than eight hours a day, having more years of work experience, and having poor social support. The study found that the odds of having DD are 7.78 times higher among those HCWs who were single as compared to those who were divorced. This result was in line with previous results from Eka kotebe hospital (29), Dessie comprehensive specialized hospital (30), and southern Ethiopia (22). Being single was the psychosocial risk factors of depressive symptoms due to a lack of a partner to express their daily stressors, thereby lacking social support. In this study, presence of childhood abuse among HCWs were 2.57 times at a higher level of depression compared to no history of child hood abuse. A systematic review and meta-analysis study also shows child maltreatment had 1.07 higher odds of having depressive disorder (36,37). The fact that effect of child hood abuse in any form leads to depression even chronic form is demonstrated by other studies that have been connected to changes in a person’s brain structure. In the worst case scenario, it can significantly raise the likelihood of adult DD and suicide (36–38).

Further, working more than 8 hour per day and having work experience of 5-15 years were associated with depression. Working more than 8 hour per day and having work experience of 5-15 years are 3 and 4 times at high risk of developing depression respectively. This is in line with studies done in Japan (39) and China (40). Long working hour leads to depression due to the fact that those who pass more of their time in work did not have enough relaxation time and those who have more year of experience have more responsibility in patient care which leads to higher risk of depression. Poor social support is another factor found to be significantly associated with depression among HCWs. Those who had poor social support had 2 times at high risk of developing depression than those who have strong social support. This is agreed with studies done in Ethiopia (41), Brazil (42) and China (43).This is due to the fact that those who had poor social support have no friends or relatives whom they can count on in time of trouble which predispose them to depression.

Furthermore, this study found that those who had a history of suicidal attempt and stressful life event ever present are significantly associated with depression. Suicidal attempt and stressful life event ever present are 2.6 and 1.52 times at high risk of developing depression respectively. This is also stated by different studies from Australia and Malaysia, in which the odds of DD increase by 1.24 among those who attempt suicidal in Australia and 55% among those who has suicidal ideation in Malaysia (44,45). Also, in the current study, back pain is another factor associated with depression among HCWs. Those HCWs who had back pain over the past 30 days are 2 times more likely to experience depressive symptoms. This result was nearly agreed by a previous studies done in Turkey in which back pain increase depressive symptoms by 61.4% (46).

### Limitation of the study

Determining causality was not possible with a cross-sectional study design. The fact that this study was limited to one institution makes it impossible to share a wealth of data regarding the prevalence of depressive symptoms among healthcare workers employed by other institution. This study’s inability to assess chronic depressive symptoms was another drawback. Furthermore, an in-depth understanding of depressed symptoms could have been attained by qualitative or mixed-method studies, but the quantitative nature of this study made this impossible.

## Conclusion

In this research, we looked at the prevalence of depressive symptoms and factors of DD among healthcare workers at SPHMMC. The outcome demonstrates that HCWs have a higher prevalence of depressed symptoms than does the rest of the population. HCWs with a history of suicidal attempt, those who had history of stressful life events, back-pain story, single, history of child hood abuse, more than eight hours of work per day, more years of work experience, and poor social support were significantly associated with depressive symptoms Therefore, this health care workers should receive special attention for early screening and treatment of depression.

## Abbreviations

COVID-19: Corona Virus Disease 2019
DASS-21: Depression Anxiety Stress Scale 21
DD: Depressive Disorder
HCWs: Health Care Workers
MDD: Major Depressive Disorder
PHQ-9: Patient Health Questionnaire-9
SPHMMC: Saint Paul’s Hospital Millennium Medical College
YLD: Years Lived with Disability

## Declarations

Ethical review board of Saint Paul’s Hospital Millennium Medical College reviewed the study protocol, and approved ethical clearance (Ref No: Pm23/651). To reach health care professionals we have got letter of permission from head of saint Paul’s hospital millennium medical college research directorate. The right of participants to discontinue or refuse participation was respected and confidentiality was maintained. Verbal informed consent was taken in all participant before taking part in this study.

## Data Availability

All relevant data are within the manuscript and its Supporting Information files

## Acknowledgment

We would like to thank the public health department at Saint Paul’s Hospital Millennium Medical College for supporting this research. We extend our heartfelt gratitude to the higher authorities at Saint Paul’s Hospital Millennium Medical College for their dedicated cooperation. We also thank all research participants for their unwavering commitment.

## Author Contributions

Conceptualization: Melese Bahiru, Berhanu Teshome, Eyoel Berhane.

Data curation: Melese Bahiru, Berhanu Teshome, Eyoel Berhane, Kidest Getu.

Formal analysis: Melese Bahiru, Berhanu Teshome.

Investigation: Melese Bahiru, Berhanu Teshome, Eyoel Berhane.

Methodology: Melese Bahiru, Berhanu Teshome,

Kidest Getu Supervision: Melese Bahiru, Kidest Getu.

Visualization: Melese Bahiru, Kidest Getu, Berhanu Teshome, Eyoel Berhane.

Writing – original draft: Melese Bahiru, Berhanu Teshome.

Writing – review & editing: Melese Bahiru, Berhanu Teshome, Eyoel Berhane.

## Notes

### Competing Interest Statement

The authors have declared no competing interest.

### Funding Statement

The author(s) received no specific funding for this work.

### Author Declarations

Ethical review board of Saint Paul’s Hospital Millennium Medical College reviewed the study protocol, and approved ethical clearance (Ref No: Pm23/651). To reach health care professionals we have got letter of permission from head of saint Paul’s hospital millennium medical college research directorate. The right of participants to discontinue or refuse participation was respected and confidentiality was maintained. Oral informed consent was taken in all participant before taking part in this study.

